# Real-world effectiveness of molnupiravir, nirmatrelvir-ritonavir, and sotrovimab on preventing hospital admission among higher-risk patients with COVID-19 in Wales: a retrospective cohort study

**DOI:** 10.1101/2023.01.24.23284916

**Authors:** Andrew Evans, Cathy Qi, Lolu Adebayo, Jonathan Underwood, James Coulson, Rowena Bailey, Gareth John, Adrian Edwards, Alison Cooper, Ronan A Lyons, Ashley Akbari

**Affiliations:** Health and Social Services Group, Welsh Government, Cathays Park, Cardiff CF10 3NQ; Population Data Science, Swansea University Medical School, Faculty of Medicine, Health and Life Science, Swansea University, Singleton Park Swansea SA2 8PP; School of Medicine, Cardiff University, University Hospital of Wales, Heath Park, Cardiff, CF14 4XN; Information Services Directorate, Digital Health and Care Wales, Tŷ Glan-yr-Afon, 21 Cowbridge Road East, Cardiff CF11 9AD; Division of Population Medicine, Director, PRIME Centre Wales and Wales COVID-19 Evidence Centre. Division of Population Medicine, Cardiff University, Neuadd Meirionnydd, Heath Park, Cardiff, CF14 4YS; Wales Covid-19 Evidence Centre. Division of Population Medicine, Cardiff University, Neuadd Meirionnydd, Heath Park, Cardiff, CF14 4YS

## Abstract

**Objective:** To compare the effectiveness of molnupiravir, nirmatrelvir-ritonavir, and sotrovimab with no treatment in preventing hospital admission or death in higher-risk patients infected with SARS-CoV-2 in the community.

**Design:** Retrospective cohort study of non-hospitalised adult patients with COVID-19 using the Secure Anonymised Information Linkage (SAIL) Databank.

**Setting:** A real-world cohort study was conducted within the SAIL Databank (a secure trusted research environment containing anonymised, individual, population-scale electronic health record (EHR) data) for the population of Wales, UK.

**Participants:** Adult patients with COVID-19 in the community, at higher risk of hospitalisation and death, testing positive for SARS-CoV-2 between 16^th^ December 2021 and 22^nd^ April 2022.

**Interventions:** Molnupiravir, nirmatrelvir-ritonavir, and sotrovimab given in the community by local health boards and the National Antiviral Service in Wales.

**Main outcome measures:** All-cause admission to hospital or death within 28 days of a positive test for SARS-CoV-2.

**Statistical analysis:** Cox proportional hazard model with treatment status (treated/untreated) as a time-dependent covariate and adjusted for age, sex, number of comorbidities, Welsh Index of Multiple Deprivation, and vaccination status. Secondary subgroup analyses were by treatment type, number of comorbidities, and before and on or after 20^th^ February 2022, when omicron BA.1 and omicron BA.2 were the dominant subvariants in Wales.

**Results:** Between 16^th^ December 2021 and 22^nd^ April 2022, 7,103 higher-risk patients were eligible for inclusion in the study. Of these, 2,040 received treatment with molnupiravir (359, 17.6%), nirmatrelvir-ritonavir (602, 29.5%), or sotrovimab (1,079, 52.9%). Patients in the treatment group were younger (mean age 53 vs 57 years), had fewer comorbidities, and a higher proportion had received four or more doses of the COVID-19 vaccine (36.3% vs 17.6%).

Within 28 days of a positive test, 628 (9.0%) patients were admitted to hospital or died (84 treated and 544 untreated). The primary analysis indicated a lower risk of hospitalisation or death at any point within 28 days in treated participants compared to those not receiving treatment. The adjusted hazard rate was 35% (95% CI: 18-49%) lower in treated than untreated participants. There was no indication of the superiority of one treatment over another and no evidence of a reduction in risk of hospitalisation or death within 28 days for patients with no or only one comorbidity. In patients treated with sotrovimab, the event rates before and on or after 20^th^ February 2022 were similar (5.0% vs 4.9%) with no significant difference in the hazard ratios for sotrovimab between the time periods.

**Conclusions:** In higher-risk adult patients in the community with COVID-19, those who received treatment with molnupiravir, nirmatrelvir-ritonavir, or sotrovimab were at lower risk of hospitalisation or death than those not receiving treatment.

## Introduction

The development of novel therapeutic agents for the treatment of SARS-CoV-2 infection has been a priority during the COVID-19 pandemic. In early 2021, the UK Government established an antiviral task force with the objective of identifying and deploying innovative COVID-19 treatments which could be taken at home to reduce disease transmission and speed up individuals’ recovery.^1^ Later in 2021, the Medicines and Healthcare products Regulatory Agency (MHRA) granted conditional marketing authorisations for three antiviral medicines, remdesivir, molnupiravir, and nirmatrelvir-ritonavir, and the two neutralising monoclonal antibody (nMAb) treatments casirivimab and imdevimab, and sotrovimab, for the treatment of mild to moderate COVID-19 in individuals with one or more risk factors for severe disease.

Whilst antiviral and nMAb therapies have been shown to reduce the risk of progression to severe disease in clinical trials, licensing studies were carried out before the deployment of COVID-19 vaccination programmes.^2-6^ Furthermore, the emergence of new SARS-CoV-2 variants shown in vitro to have the ability to evade neutralisation by monoclonal antibodies,^7^ has cast doubts on the sustained effectiveness of these treatments with international medicines regulators and the manufacturer restricting the use of casirivimab and imdevimab,^8,9^ and the World Health Organisation (WHO) now strongly recommending against the use of sotrovimab in patients with non-severe COVID-19.^10^

There is therefore considerable uncertainty as to whether the benefits of treatments observed in a clinical trial are realised in the real world in highly vaccinated populations and where newer variants dominate those that resulted in infection amongst trial participants. To address these concerns, in the UK, the deployment of antiviral and nMAb treatments to non-hospitalised patients testing positive for COVID-19 was restricted to those in tightly defined cohorts whose immune systems mean they remain at higher-risk of serious illness, despite vaccination.^11^

The UK deployment of COVID-19 treatments to non-hospitalised higher-risk patients began on 16^th^ December 2021. A clinical access policy continues to support the treatment of higher-risk patients with antiviral medicines, and in limited circumstances, sotrovimab^12^ and over 96,000 people in England and Wales have subsequently received treatment.^13,14^ Further real-world evidence of the effectiveness of the UK deployment approach is urgently needed.

In this retrospective cohort study, we sought to compare the effectiveness of sotrovimab, molnupiravir, and nirmatrelvir-ritonavir in preventing hospital admission and death in higher-risk patients (Figure 1) with COVID-19 in Wales, UK during the first five months of deployment, using anonymised, individual-level, population-scale electronic health record (EHR) data in the Secure Anonymised Information Linkage (SAIL) Databank^15-19^ trusted research environment (TRE), accounting for age, comorbidity, socioeconomic deprivation, and vaccination status. We also conducted a subgroup analysis of patients treated with sotrovimab before and following the emergence of the omicron BA.2 variant in Wales.^20^

**Figure 1:**
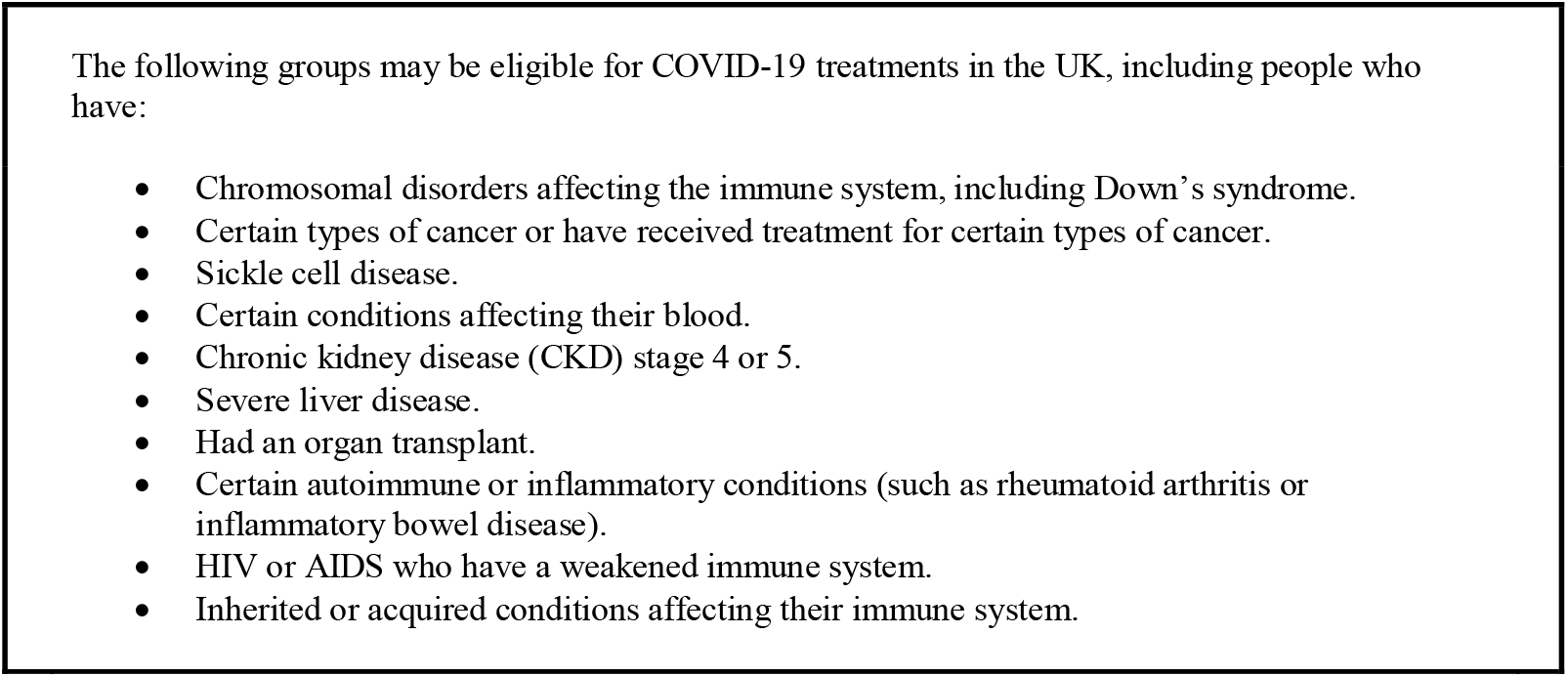
Higher-risk patients eligible for COVID-19 treatments

## Methods

### Study Design and Population

Retrospective cohort study in Wales of non-hospitalised adult patients with COVID-19. To be eligible for inclusion in the study, participants needed to have a positive SARS-CoV-2 polymerase chain reaction (PCR) or lateral flow device (LFD) test between 16^th^ December 2021 and 22^nd^ April 2022, and be included in one or more of the ten cohorts considered to be at higher-risk from COVID-19 in accordance with the UK clinical access policy, and who were eligible for treatment with sotrovimab, molnupiravir, or nirmatrelvir–ritonavir.^11^ Participants were retrospectively followed up for 28 days following the index date (the date of positive PCR or LFD test) for any cause hospitalisation or death.

### Data sources and variables

Anonymised individual-level, population-scale, linked, routinely-collected electronic health record (EHR) data within the SAIL Databank were used. Eligible participants were identified by Digital Health and Care Wales (DHCW) from hospital episode data contained within the Patient Episode Database for Wales (PEDW), and primary care prescribing data linked to PCR and LFD test results in the Welsh Laboratory Information System (WLIMS), or from opportunistic referral by clinicians. Data available included the date of a positive test, participants’ Lower-level Super Output Area (LSOA) of residence (administrative authority locality), and information about the clinical condition conferring eligibility for each participant.

An individual-level dataset of patients treated with antiviral and nMAb treatments was obtained from each of Wales’ seven local health boards (LHBs) and the National Antiviral Service (NAVS)^21^ and linked with the cohort of eligible people identified as described above. People hospitalised on the day of the positive test, who received treatment but who had no record of positive PCR or LFD test either in the data provided by DHCW or contained within the SAIL Databank, people who received a study treatment before their index date, people hospitalised or who died before or on the index date, and anyone treated more than seven days after the most recent positive test, were excluded. Further exclusions were applied to individuals still missing key demographic information (age, sex, LSOA) or who had a non-Welsh LSOA.

### Exposure

The exposure was treatment with molnupiravir, nirmatrelvir-ritonavir, or sotrovimab. Participants were either treated within seven days of a positive PCR or LFD test (days 0 to 7) or were untreated with one of the treatments under investigation within 28 days of a positive test.

### Outcomes

The primary outcome was any cause hospitalisation or death (if death occurred without prior admission) within 28 days of a positive PCR or LFD test. Participants with no record of a hospital admission within 28 days were assumed to be not hospitalised, and those with no record of death were assumed to be alive.

### Covariates

Participants’ baseline covariates included age, sex, number of comorbidities, Charlson comorbidity index (CCI) score, clinical subgroup (categorised as immunosuppressed conditions including haematological cancers, non-haematological cancers, other high-risk conditions, or unknown), Welsh Index of Multiple Deprivation (WIMD) version 2019 as quintiles mapped from LSOAs, COVID-19 vaccination status (unvaccinated, one to three vaccinations, or four or more vaccinations), and type of treatment received (molnupiravir, nirmatrelvir-ritonavir, or sotrovimab).

### Statistical Analysis

Kaplan-Meier graphs were used to show event-free survival during the 28 day observation period. Event-free survival was presented by demographic factors (sex, age, WIMD), clinical factors (vaccination, number of comorbidities, weighted CCI score, clinical subgroup), and treatment groups.

In the primary analysis, the risk of hospitalisation or death from the index date to day 28 was analysed using a Cox proportional hazard model with treatment status (treated/untreated) as a time-dependent covariate and adjusted for age, sex, number of comorbidities, WIMD, and vaccination status. For those receiving treatment, treatment status was updated from untreated to treated the day after treatment if they remained in the risk set. Time-to-event was from the index date and censored at 28 days for those without an event by day 28. The proportional hazards assumption was assessed using Schoenfeld’s global test and visual plots.

To assess the sensitivity of the primary result to possible bias, we repeated the analysis not adjusting for confounders, adjusting for the CCI weighted score rather than the number of comorbidities, and adjusting for the clinical subgroup instead of the number of comorbidities. We repeated the primary model without the time-dependent component, comparing two fixed groups of individuals (treated and untreated) and included all individuals who received treatment (at any time point) in the treated group. Finally we performed a logistic regression analysis with all-cause hospitalisation or death as a binary outcome comparing treated and untreated groups (with all individuals who received treatment in the treated group), and adjusting for the covariates included in the primary model. Hazard ratios (HR) and odds ratios (OR) were calculated with 95% confidence intervals (CIs).

Secondary analyses assessed the primary outcome by treatment type, number of comorbidities, and before and on or after 20^th^ February 2022.

We estimated the HR (95% CI) associated with each treatment type by fitting the primary model with treatment status categorised into molnupiravir, nirmatrelvir-ritonavir, sotrovimab, or untreated. The HR associated with being currently treated vs untreated was estimated for each comorbidity category by the addition of an interaction term between comorbidity and treatment status in the primary model. The analysis for before or on or after 20^th^ February 2022 included a subset of participants who received sotrovimab or were untreated, and the HR (95% CI) associated with treatment with sotrovimab was estimated for each time point by addition of time point (before or on or after 20^th^ February 2022) and an interaction term between time point and treatment status in the primary model.

All statistical analyses were conducted using R V4.1.3 and STATA 17.0 (StataCorp. 2021. Stata Statistical Software: Release 17. College Station, TX: StataCorp LLC.)

## Results

We identified 10825 high-risk individuals, of whom 7,128 tested positive between 16^th^ December 2021 and 22^nd^ April 2022. A further 603 individuals not identified by DHCW were identified from the LHB and NAVS treatment dataset. In total 7,013 patients with a positive PCR or LFD test were considered to meet the clinical eligibility criteria for antiviral or nMAb treatment (Figure 2). Of these, 2,040 (29.1%) received treatment within seven days of a positive test and were not admitted to hospital on or before receiving treatment, 32 (0.5%) were hospitalised within seven days of a positive test and received treatment on the day or after the day of admission, and 4,941 (70.5%) did not receive treatment in the community within the study period.

**Figure 2:**
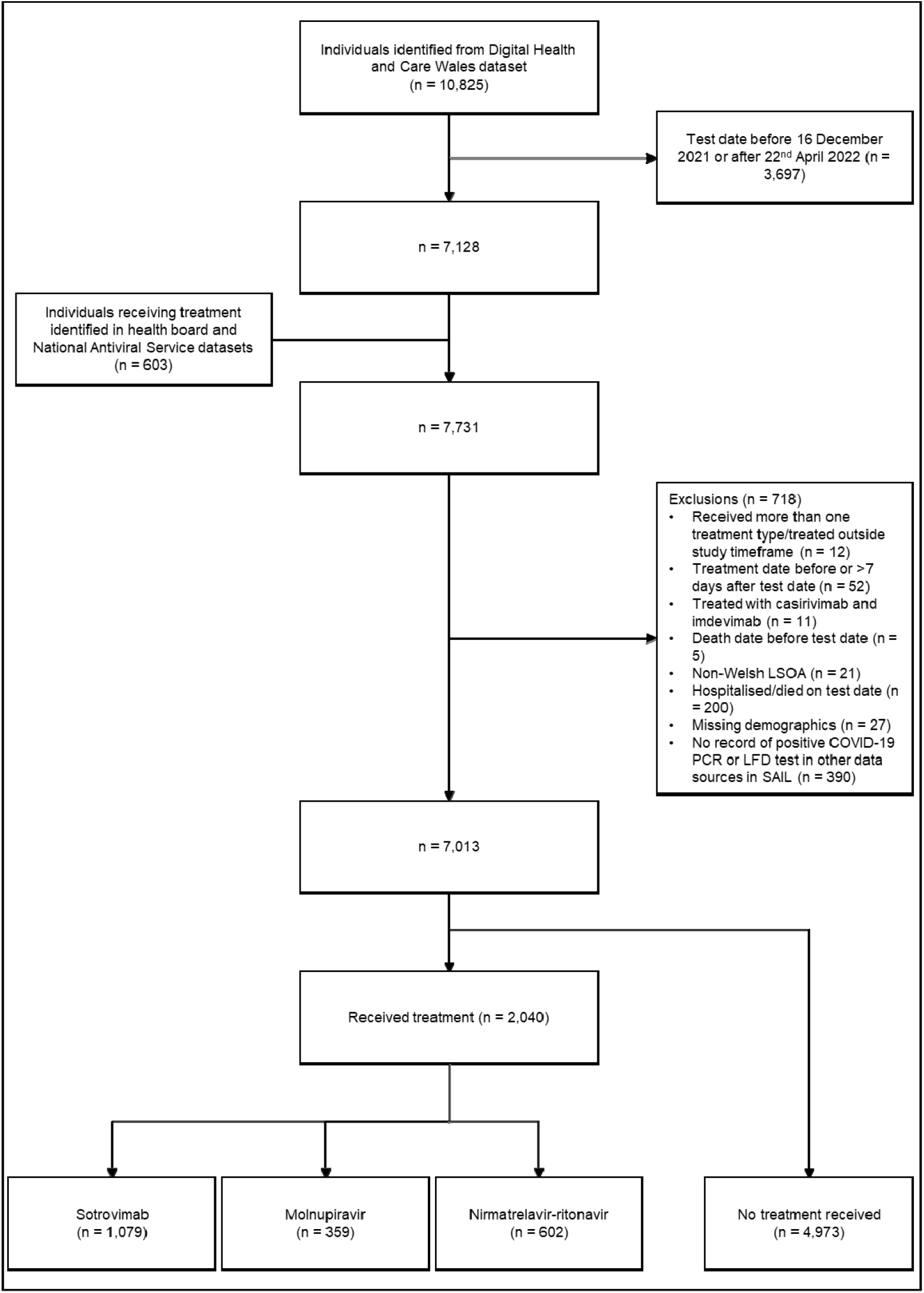
Study participant flowchart

The baseline characteristics of the study population are presented in Table 1. Patients in the treatment group were younger (mean age 53 vs 57 years), had fewer comorbidities, and a higher proportion had received four or more doses of COVID-19 vaccine (36.3% vs 17.6%).

**Table 1:**
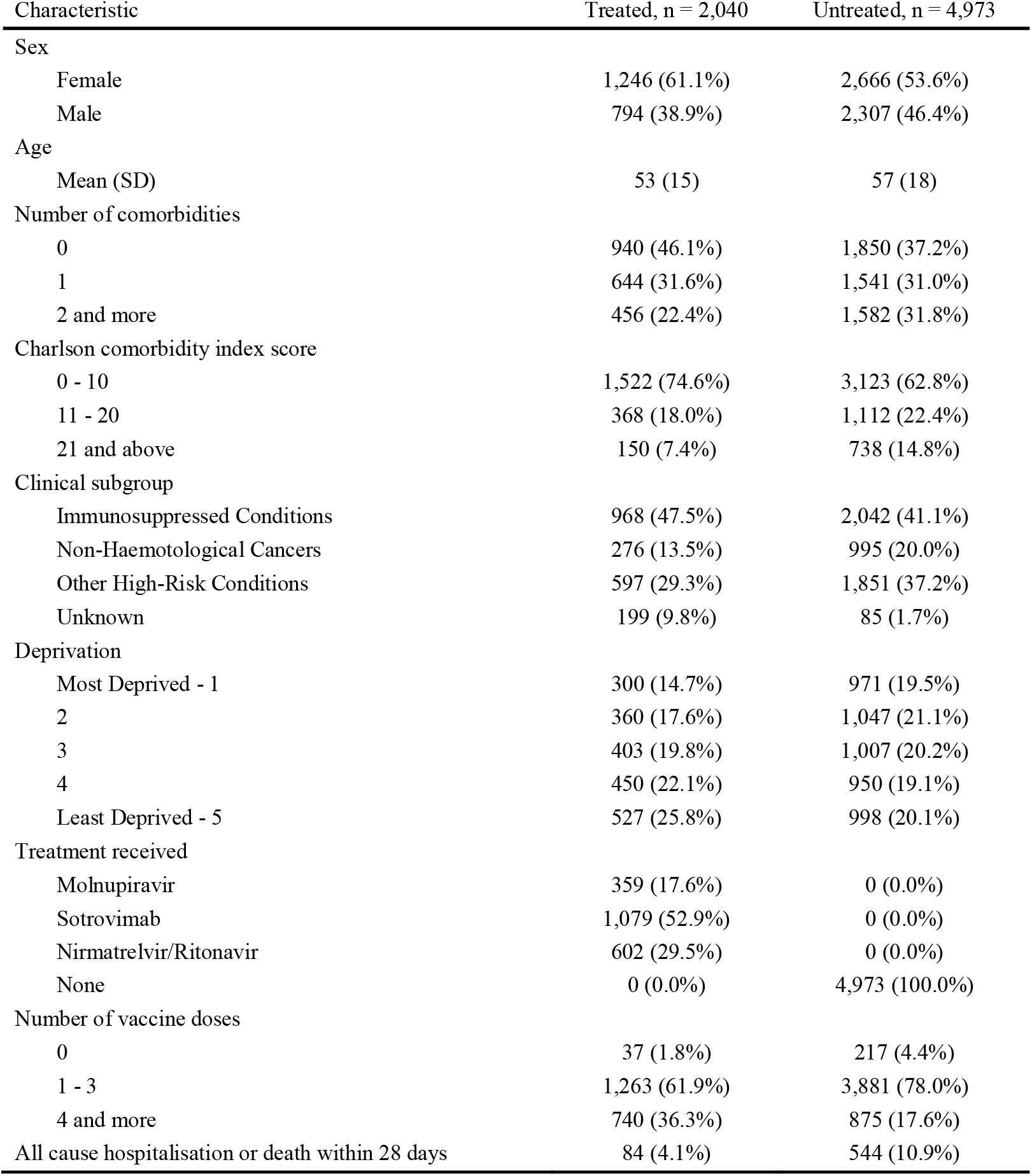
Baseline characteristics of study participants

### Association between demographic and clinical characteristics and the primary outcome

The probability of avoiding hospital admission or death within 28 days was higher in younger age groups (aged under 60), amongst patients living in the lowest quintile of multiple deprivation (least deprived quintile), and in those who had received four or more doses of COVID-19 vaccine when compared to those receiving fewer doses. Those receiving any COVID-19 vaccinations had a lower probability of admission or death than unvaccinated patients. Notable differences were observed in event-free survival between those with comorbidities (when measured either by the number or by the CCI score) with lesser differentiation observed between the clinical subgroups (Figure 3).

**Figure 3:**
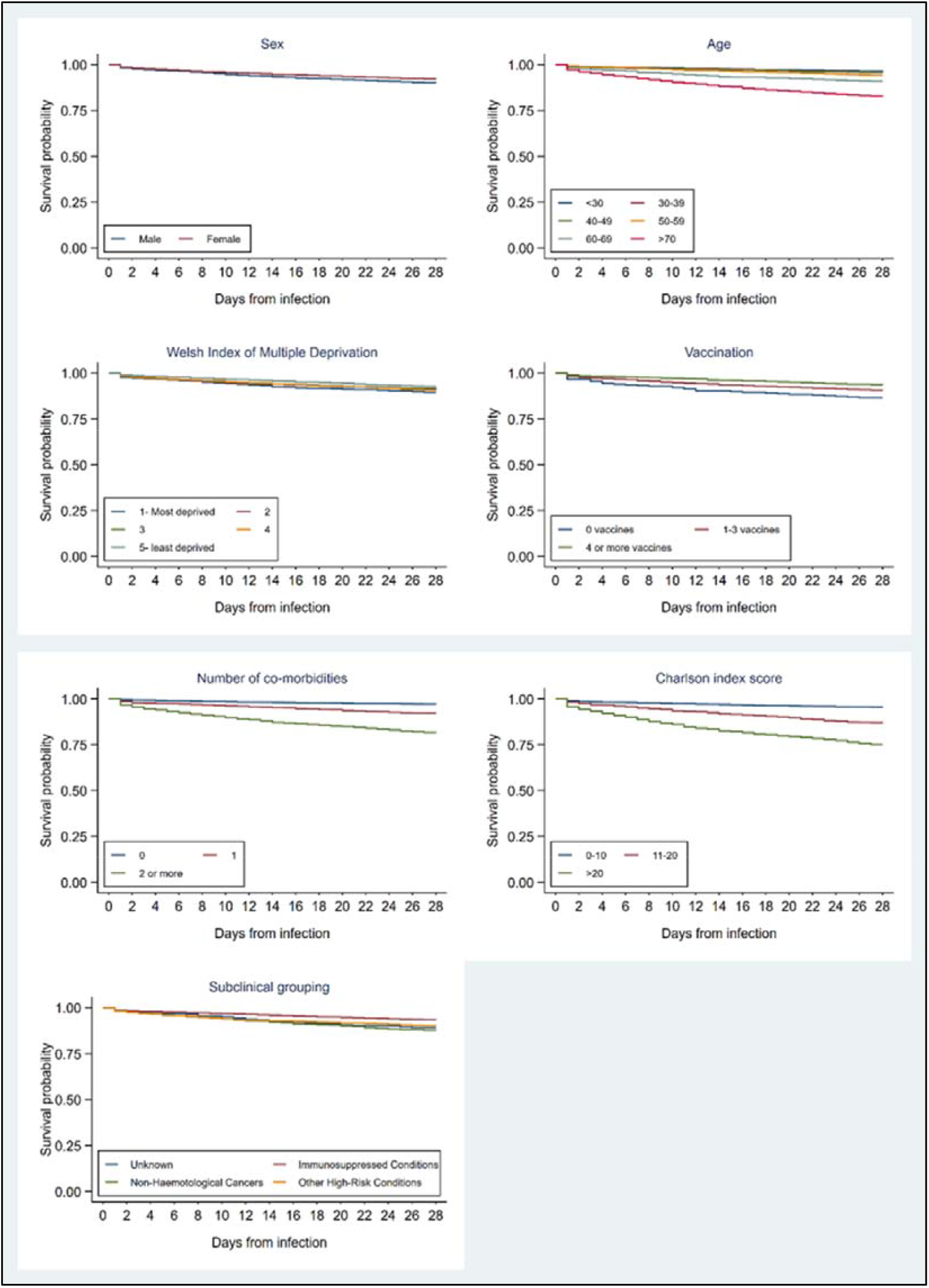
Kaplan-Meier charts showing association between demographic and clinical factors with event-free survival during the 28-day observation period following the date of COVID-19 infection

### Treatment effectiveness for the primary outcome

In total 628 (9.0%) hospitalisations or deaths within 28 days of a positive test were observed in the study period; 84 (4.1%) in treated and 544 (10.9%) in untreated participants.

The primary analysis results indicated a lower risk of hospitalisation or death at any point within 28 days in treated participants compared to those not receiving treatment. The estimated hazard rate was 35% (95% CI: 18-49%) lower in treated than untreated participants after adjusting for confounders and 52% (95% CI: 39-62%) lower in the unadjusted analysis. The results of the primary and sensitivity analyses are shown in Table 2.

**Table 2:** Primary outcome and sensitivity analyses

|  | Treated | Untreated |
| --- | --- | --- |
| Primary analysis: (adjusted for age, sex, WIMD, vaccination status, number of comorbidities) |  |  |
| Numbers included | 2,040 | 4,973 |
| No. of events (%) | 84 (4.1) | 544 (10.9) |
| Hazard ratio (95% CI) | 0.65 (0.51- 0.82) |  |
| Sensitivity analysis: unadjusted |  |  |
| Numbers included | 2,040 | 4,973 |
| No. of events (%) | 84 (4.1) | 544 (10.9) |
| Hazard ratio (95% CI) | 0.48 (0.38- 0.61) |  |
| Sensitivity analysis: (adjusted for age, sex, WIMD, vaccination status, clinical group) |  |  |
| Numbers included | 2,040 | 4,973 |
| No. of events (%) | 84 (4.1) | 544 (10.9) |
| Hazard ratio (95% CI) | 0.61 (0.48- 0.78) |  |
| Sensitivity analysis: (adjusted for age, sex, WIMD, vaccination status, weighted CCI score) |  |  |
| Numbers included | 2,040 | 4,973 |
| No. of events (%) | 84 (4.1) | 544 (10.9) |
| Hazard ratio (95% CI) | 0.67 (0.53- 0.85) |  |
| Sensitivity analysis: time-independent (adjusted for age, sex, WIMD, vaccination status, number of comorbidities) |  |  |
| Numbers included | 2,072 | 4,941 |
| No. of events (%) | 116 (5.6) | 512 (10.4) |
| Hazard ratio (95% CI) | 0.72 (0.58- 0.88) |  |
| Sensitivity analysis: logistic model (adjusted for age, sex, WIMD, vaccination status, number of comorbidities)<sup>1</sup> |  |  |
| Numbers included | 2,072 | 4,941 |
| No. of events (%) | 116 (5.6) | 512 (10.4) |
| Odds ratio (95% CI) | 0.70 (0.57- 0.88) |  |

1. The treated group included 32 individuals who received treatment after hospitalisation

### Secondary analyses

Of the 2,040 patients receiving any treatment, 359 (17.6%) received molnupiravir, 602 (29.5%) nirmatrelvir-ritonavir, and 1,079 (52.9%) sotrovimab. The event rates were 3.9% (14/359) for molnupiravir, 2.8% (17/602) for nirmatrelvir-ritonavir, and 4.9% (53/1,079) for sotrovimab. Each treatment was found to lower the risk of hospitalisation or death when compared to no treatment. The adjusted HRs for patients treated with molnupiravir, nirmatrelvir-ritonavir, and sotrovimab were 0.49 (95%CI: 0.29-0.83), 0.59 (95%CI: 0.36-0.97), and 0.73 (95%CI: 0.55-0.98) respectively. We found no indication of the superiority of one treatment over another.

When we examined the effect of comorbidity on treatment outcome, there was no evidence of a reduction in risk of hospitalisation or death within 28 days for patients with no or only one comorbidity. For patients with two or more comorbidities the adjusted HR was 0.45 (95%CI: 0.31-0.65) indicating a 55% reduction in hazard after treatment (Table 3).

**Table 3:** Secondary analysis: subgroup effect (number of comorbidities)

|  | Treated, n = 2,040 | Untreated, n = 4,973 |
| --- | --- | --- |
| Numbers included (%) |  |  |
| 0 | 940 (46.1) | 1,850 (37.2) |
| 1 | 644 (31.6) | 1,541 (31.0) |
| ≥ 2 | 456 (22.4) | 1,582 (31.8) |
| No. of events (%) |  |  |
| 0 | 22 (2.3) | 58 (3.1) |
| 1 | 31 (4.8) | 140 (9.1) |
| ≥ 2 | 31 (6.8) | 346 (21.9) |
| Hazard ratio (95% CI) |  |  |
| 0 | 1.09 (0.67- 1.79) |  |
| 1 | 0.79 (0.53- 1.17) |  |
| ≥ 2 | 0.45 (0.31- 0.65) |  |

The subgroup analysis including 6,052 participants (1079 patients treated with sotrovimab and 4973 not treated) showed 461 (42.7%) treated before and 618 (57.3%) treated on or after 20^th^ February 2022. The number of events occurring before and on or after this date were 23 and 30 respectively, and the event rates were similar (5.0% vs 4.9%). No significant difference was observed in the HRs for sotrovimab between the time periods when omicron BA.1 and omicron BA.2 were the dominant subvariants in Wales (Table 4).

**Table 4:** Secondary analysis: subgroup before and on or after 20^th^ February 2022

|  | Treated, n= 1,079 | Untreated, n= 4,973 |
| --- | --- | --- |
| Numbers included (%) |  |  |
| Before | 461 (42.7) | 2,457(49.4) |
| After | 618 (57.3) | 2,516 (50.6) |
| No. of events (%) |  |  |
| Before | 23 (5.0) | 269 (11.0) |
| After | 30 (4.9) | 275 (10.9) |
| Hazard ratio (95% CI) |  |  |
| Before | 0.76 (0.50- 1.18) |  |
| After | 0.70 (0.48- 1.03) |  |

The baseline characteristics of patients included in the secondary analyses are available in Supplementary Tables S1 – S3).

## Discussion

In this retrospective real-world cohort study of highest-risk, non-hospitalised patients with COVID-19, prompt treatment with molnupiravir, nirmatrelvir-ritonavir, or sotrovimab was associated with significantly lower risks of all-cause hospital admission or death within 28 days of testing positive for SARS-CoV-2.

To our knowledge this is the first real-world study to compare the targeted deployment of antiviral and nMAb treatment to only patients considered to be at the very highest risk of severe disease and the effectiveness of the deployment model being used in the UK against untreated patients in similar high-risk cohorts. In the main analysis, we found untreated patients considered to be at the highest-risk continue to face a substantial risk of hospitalisation or death when they had COVID-19 and whilst not eliminated, that after controlling for a wide range of potential confounders, that risk was significantly reduced by treatment with one of molnupiravir, nirmatrelvir-ritonavir, or sotrovimab. In subgroup analysis we found no clear evidence for any one treatment over any other. Secondary analyses suggest the greatest benefits of treatment are for patients with multiple comorbidities, and that there was little change to the effectiveness of sotrovimab following the emergence of the omicron BA.2 subvariant.

### Findings in context

We found people in the 10 high-risk cohorts eligible in the UK continue to face a substantial (10.9%) risk of hospitalisation or death when they have COVID-19. The reduction in hospitalisations and deaths found in this study are broadly consistent with published pre-omicron randomised controlled trials for sotrovimab,^2^ molnupiravir,^3^ and nirmatrelvir-ritonavir,^4^ despite our study being carried out when omicron was the predominant variant in Wales. Similar results were observed in real-world studies of nirmatrelvir-ritonavir^22^ where a significant decrease in the rate of severe COVID-19 or death was observed with an adjusted HR of 0.54 (95%CI: 0.39-0.75) when omicron was the predominant variant, a preprint study of sotrovimab with a 55% relative risk (RR) reduction of hospitalisation (RR: 0.45, 95%CI: 0.41-0.49) before^23^ and a study of sotrovimab with a 72% reduction in risk of hospitalisation (OR: 0.28, 95%CI: 0.11-0.71) after the emergence of omicron variants.^24^

Our results contrast to those of the recently published 25,000 participant, prospective, open-label, UK-wide, PANORAMIC trial, which found only a 1% risk of all-cause hospitalisation and that molnupiravir did not reduce the frequency of COVID-19-associated hospitalisations or death among adults over 50 years of age or over 18 with other risk factors, in the community.^25^

The populations included in these studies have major differences to those in our analysis and included unvaccinated patients^2-4^ and individuals from relatively lower-risk cohorts, ^22-25^ meaning their findings are unlikely to be generalisable to the current highly targeted deployment in the UK.

Few studies have been limited to comparable higher-risk populations. Those which have studied similar cohorts, including one peer-reviewed and one pre-print study exploring the UK’s targeted deployment, also reported lower rates of hospital admission and death amongst those receiving treatment.^26,27^ Whilst in contrast to the findings of our secondary analysis, Zheny et al reported sotrovimab treatment was associated with a reduced risk of death or hospitalisation within 28 days of a positive COVID-19 test when compared with molnupiravir, neither that study nor the pre-print study conducted by Patel et al, were designed to compare the effectiveness of treatment to no treatment, in the high-risk cohort.

Only one small study limited to solid organ transplant recipients (who are included amongst the high-risk groups eligible for treatment in the UK), compared treatment with sotrovimab, molnupiravir, or nirmatrelvir-ritonavir to no treatment and suggested a 13% absolute reduction in hospital admissions at 30 days (14% vs 27%) amongst those receiving treatment. As in our study, no difference was observed between treatments.^28^

Despite conflicting evidence regarding sotrovimab’s possible loss of efficacy against omicron BA.2 and subsequent subvariants, we found no difference between the efficacy of sotrovimab before or during the period when omicron BA.2 was the predominant variant in Wales, suggesting a continued protective effect of sotrovimab against this subvariant. This finding was similar to the exploratory analysis undertaken by Zheny et al, and evidence that sotrovimab is capable of neutralising omicron subvariants BA.2, BA.2.12.1, BA.4 and BA.5 in vitro at concentrations 47-fold lower than the maximum plasma concentration and 10-fold lower than the mean 28-day plasma concentration.^29^

### Policy implications

The most recent iteration of the UK clinical access policy^12^ places nirmatrelvir-ritonavir as the first-line treatment option, followed by remdesivir and molnupiravir, with sotrovimab now reserved for exceptional cases where antiviral treatments are contraindicated or unsuitable. Contraindications to the use of nirmatrelavir-ritonavir include drug-drug interactions, which can lead to serious or life-threatening drug toxicities. There are practical challenges with the administration of three-day courses of intravenous remdesivir to non-hospitalised patients; and concerns about the effectiveness of molnupiravir^30^ and sotrovimab^10^ could result in some people at high-risk not receiving treatment. As superiority of one treatment over others was not evident in our results, we argue for continued access to all treatments within the clinical access policy. Our findings support the continuation of the UK’s policy to target antiviral and nMAb therapy deployment to those at the highest risk.

Results of subgroup analyses suggested only patients with multiple comorbidities had a reduced risk of hospitalisation, which would support further prioritisation of treatment within the highest-risk cohort.

### Strengths and weaknesses

The key strengths of this study are its relatively large size and the completeness of the data sources available within the SAIL Databank. Access to the positive PCR or LFD test results for all people in the eligible cohort allowed treatment to be compared to no treatment in similar groups to assess the effectiveness of providing treatment rather than between treatments. The concurrent national deployment of molnupiravir and sotrovimab, and subsequently nirmatrelvir-ritonavir, allowed direct comparison between treatments. Finally, the duration of the study allowed a comparison of the effectiveness of sotrovimab before and following the emergence of the omicron BA.2 subvariant.

There are several limitations of the study. We cannot discount the possibility of selection bias given the observational design. The 10 groups of high-risk patients are heterogeneous and whilst we controlled for three clinical subgroups and comorbidity, it is plausible that differences between individuals in the treatment and control groups persist. For example people in the treatment group were generally younger, had fewer comordibities and were more likely to be fully vaccinated but might have been considered more unwell than those not treated. Indeed asymptomatic patients were not eligible for treatment under the current UK clinical access policyand unlike the treatment group, the control group did not exclude any patients who would have been found not to be in one of the 10 high-risk cohorts on clinical screening. Therefore whilst the treated group had some characteristics which might reduce the likelihood of hospitalisaton, over-representation of less unwell and lower-risk patients were in the control group could also lead an underestimate oftreatment benefits. We included all cause rather than COVID-19 related, hospitalisation and deaths within 28 days of a positive COVID-19 test. Thus we have not discounted non-COVID-19 causes of admission which may be more prevalent in the treatment group due to possible differences in the characteristics of people in treatment and non-treatment groups. Nevertheless, we believe this reflects the changing pattern of COVID-19 in the UK where admissions where COVID-19 was the primary cause have declined since early 2022.^31^

Findings in observational studies should be interpreted with caution, however we observed a large effect size after adjusting for several potential confounders, which was confirmed by multiple sensitivity analyses; any bias would need to be considerable to completely account for our findings.

### Further research

Reducing unplanned hospitalisations remains a priority for health services in all parts of the UK, and there appears to be a significant beneficial effect on admissions from providing treatment to high-risk groups in the community. However, the treatments studied are not inexpensive: treatment courses of sotrovimab and nirmatrelvir-ritonavir have UK list prices of £2,209 and £829 respectively^32^ and whilst the UK price is not publicly available, the US price of a treatment course of molnupiravir has been reported as $707.^33^ The National Institute for Health and Care Excellence (NICE) has plans to publish guidance on the cost-effectiveness of these and other COVID-19 treatments in 2023.^32^ Given the limited number of real-world studies generalisable to the UK, there is a clear need to determine the relative clinical effectiveness and cost-effectiveness of different treatment options taking account of deployment as well as acquisition costs.

### Conclusion

This retrospective cohort study of non-hospitalised high-risk patients with COVID-19 suggests that prompt treatment with the oral antiviral medicines molnupiravir or nirmatrelvir-ritonavir, or the nMAb sotrovimab, was associated with a significant reduction in all-cause hospitalisation and death within 28 days of infection immediately before and during a pandemic wave in which the SARS-CoV-2 omicron BA.2 subvariant was dominant. Our findings support the UK deployment approach and the continued use of oral antiviral medicines and sotrovimab in this population, and contribute evidence to the ongoing debate on the real-world effectiveness of sotrovimab.

## Data Availability

The anonymised individual-level data sources used in this study are available in the SAIL Databank at Swansea University, Swansea, UK, but as restrictions apply, they are not publicly available. All proposals to use SAIL data are subject to review by the independent Information Governance Review Panel (IGRP). Before any data can be accessed, approval must be given by the IGRP. The IGRP gives careful consideration to each project to ensure proper and appropriate use of SAIL data. When access has been granted, it is gained through a privacy-protecting safe haven and remote access system referred to as the SAIL Gateway. SAIL has established an application process to be followed by anyone who would like to access data via SAIL at: https://www.saildatabank.com/application-process/

---

**Supplementary Table S1:** Demographic and clinical characteristics, and event rates by treatment group for individuals with 0, 1, ≥ 2 conditions on the CCI

| Characteristic | 0 conditions | | 1 condition | | $\geq 2$ conditions | |
| --- | --- | --- | --- | --- | --- | --- |
|  | Treated, n = 940 | Untreated, n = 1,850 | Treated, n = 644 | Untreated, n = 1,541 | Treated, n = 456 | Untreated, n = 1,582 |
| Sex |  |  |  |  |  |  |
| Female | 611 (65.0%) | 1,029 (55.6%) | 402 (62.4%) | 879 (57.0%) | 233 (51.1%) | 758 (47.9%) |
| Male | 329 (35.0%) | 821 (44.4%) | 242 (37.6%) | 662 (43.0%) | 223 (48.9%) | 824 (52.1%) |
| Age |  |  |  |  |  |  |
| Mean (SD) | 47 (14) | 47 (16) | 55 (15) | 58 (16) | 62 (15) | 69 (15) |
| Charlson comorbidity index score |  |  |  |  |  |  |
| 0 - 10 | 940 (100.0%) | 1,850 (100.0%) | 514 (79.8%) | 1,126 (73.1%) | 68 (14.9%) | 147 (9.3%) |
| 11 - 20 | 0 (0.0%) | 0 (0.0%) | 130 (20.2%) | 415 (26.9%) | 238 (52.2%) | 697 (44.1%) |
| 21 and above | 0 (0.0%) | 0 (0.0%) | (0.0%) | (0.0%) | 150 (32.9%) | 738 (46.6%) |
| Clinical subgroup |  |  |  |  |  |  |
| Immunosuppressed Conditions | 540 (57.4%) | 1,084 (58.6%) | 276 (42.9%) | 567 (36.8%) | 152 (33.3%) | 391 (24.7%) |
| Non-Haematological Cancers | 13 (1.4%) | 39 (2.1%) | 175 (27.2%) | 535 (34.7%) | 88 (19.3%) | 421 (26.6%) |
| Other High-Risk Conditions | 305 (32.4%) | 717 (38.8%) | 122 (18.9%) | 393 (25.5%) | 170 (37.3%) | 741 (46.8%) |
| Unknown | 82 (8.7%) | 10 (0.5%) | 71 (11.0%) | 46 (3.0%) | 46 (10.1%) | 29 (1.8%) |
| Deprivation |  |  |  |  |  |  |
| Most Deprived - 1 | 117 (12.4%) | 350 (18.9%) | 98 (15.2%) | 277 (18.0%) | 85 (18.6%) | 344 (21.7%) |
| 2 | 168 (17.9%) | 393 (21.2%) | 108 (16.8%) | 310 (20.1%) | 84 (18.4%) | 344 (21.7%) |
| 3 | 184 (19.6%) | 371 (20.1%) | 114 (17.7%) | 317 (20.6%) | 105 (23.0%) | 319 (20.2%) |
| 4 | 211 (22.4%) | 369 (19.9%) | 146 (22.7%) | 280 (18.2%) | 93 (20.4%) | 301 (19.0%) |
| Least Deprived - 5 | 260 (27.7%) | 367 (19.8%) | 178 (27.6%) | 357 (23.2%) | 89 (19.5%) | 274 (17.3%) |
| Treatment received |  |  |  |  |  |  |
| Molnupiravir | 144 (15.3%) | 0 (0.0%) | 107 (16.6%) | 0 (0.0%) | 108 (23.7%) | 0 (0.0%) |
| Sotrovimab | 447 (47.6%) | 0 (0.0%) | 347 (53.9%) | 0 (0.0%) | 285 (62.5%) | 0 (0.0%) |
| Nirmatrelvir/Ritonavir | 349 (37.1%) | 0 (0.0%) | 190 (29.5%) | 0 (0.0%) | 63 (13.8%) | 0 (0.0%) |
| None | 0 (0.0%) | 1,850 (100.0%) | 0 (0.0%) | 1,541 (100.0%) | 0 (0.0%) | 1,582 (100.0%) |
| Number of vaccine doses |  |  |  |  |  |  |
| 0 | 13 (1.4%) | 98 (5.3%) | 16 (2.5%) | 71 (4.6%) | 8 (1.8%) | 48 (3.0%) |
| 1 - 3 | 599 (63.7%) | 1,429 (77.2%) | 367 (57.0%) | 1,178 (76.4%) | 297 (65.1%) | 1,274 (80.5%) |
| 4 and more | 328 (34.9%) | 323 (17.5%) | 261 (40.5%) | 292 (18.9%) | 151 (33.1%) | 260 (16.4%) |
| All cause hospitalisation or death within 28 days | 22 (2.3%) | 58 (3.1%) | 31 (4.8%) | 140 (9.1%) | 31 (6.8%) | 346 (21.9%) |

**Supplementary Table S2:**
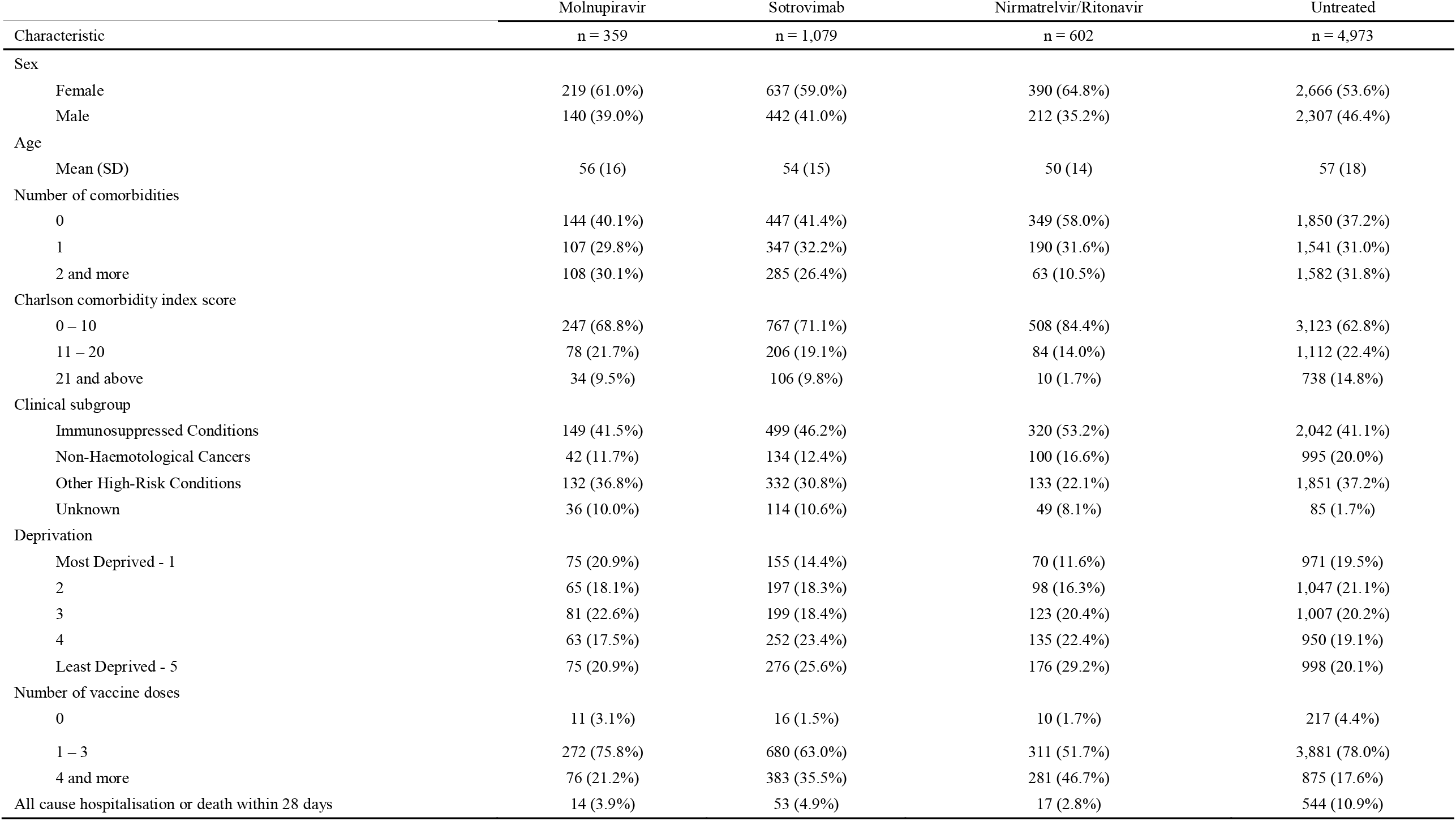
Demographic and clinical characteristics, and event rates by treatment group for each of the study treatments

|  | Molnupiravir | Sotrovimab | Nirmatrelvir/Ritonavir | Untreated |
| --- | --- | --- | --- | --- |
| Characteristic | n = 359 | n = 1,079 | n = 602 | n = 4,973 |
| Sex |  |  |  |  |
| Female | 219 (61.0%) | 637 (59.0%) | 390 (64.8%) | 2,666 (53.6%) |
| Male | 140 (39.0%) | 442 (41.0%) | 212 (35.2%) | 2,307 (46.4%) |
| Age |  |  |  |  |
| Mean (SD) | 56 (16) | 54 (15) | 50 (14) | 57 (18) |
| Number of comorbidities |  |  |  |  |
| 0 | 144 (40.1%) | 447 (41.4%) | 349 (58.0%) | 1,850 (37.2%) |
| 1 | 107 (29.8%) | 347 (32.2%) | 190 (31.6%) | 1,541 (31.0%) |
| 2 and more | 108 (30.1%) | 285 (26.4%) | 63 (10.5%) | 1,582 (31.8%) |
| Charlson comorbidity index score |  |  |  |  |
| 0 – 10 | 247 (68.8%) | 767 (71.1%) | 508 (84.4%) | 3,123 (62.8%) |
| 11 – 20 | 78 (21.7%) | 206 (19.1%) | 84 (14.0%) | 1,112 (22.4%) |
| 21 and above | 34 (9.5%) | 106 (9.8%) | 10 (1.7%) | 738 (14.8%) |
| Clinical subgroup |  |  |  |  |
| Immunosuppressed Conditions | 149 (41.5%) | 499 (46.2%) | 320 (53.2%) | 2,042 (41.1%) |
| Non-Haematological Cancers | 42 (11.7%) | 134 (12.4%) | 100 (16.6%) | 995 (20.0%) |
| Other High-Risk Conditions | 132 (36.8%) | 332 (30.8%) | 133 (22.1%) | 1,851 (37.2%) |
| Unknown | 36 (10.0%) | 114 (10.6%) | 49 (8.1%) | 85 (1.7%) |
| Deprivation |  |  |  |  |
| Most Deprived - 1 | 75 (20.9%) | 155 (14.4%) | 70 (11.6%) | 971 (19.5%) |
| 2 | 65 (18.1%) | 197 (18.3%) | 98 (16.3%) | 1,047 (21.1%) |
| 3 | 81 (22.6%) | 199 (18.4%) | 123 (20.4%) | 1,007 (20.2%) |
| 4 | 63 (17.5%) | 252 (23.4%) | 135 (22.4%) | 950 (19.1%) |
| Least Deprived - 5 | 75 (20.9%) | 276 (25.6%) | 176 (29.2%) | 998 (20.1%) |
| Number of vaccine doses |  |  |  |  |
| 0 | 11 (3.1%) | 16 (1.5%) | 10 (1.7%) | 217 (4.4%) |
| 1 – 3 | 272 (75.8%) | 680 (63.0%) | 311 (51.7%) | 3,881 (78.0%) |
| 4 and more | 76 (21.2%) | 383 (35.5%) | 281 (46.7%) | 875 (17.6%) |
| All cause hospitalisation or death within 28 days | 14 (3.9%) | 53 (4.9%) | 17 (2.8%) | 544 (10.9%) |

**Supplementary Table S3:** Demographic and clinical characteristics, and event rates by treatment group for each of the study treatments

| Characteristic | Treated before 20<sup>th</sup> February 2022 |  | Treated on or after 20<sup>th</sup> February 2022 |  |
| --- | --- | --- | --- | --- |
|  | Treated, n = 461 | Untreated, n = 2,457 | Treated, n = 618 | Untreated, n = 2,516 |
| Sex |  |  |  |  |
| Female | 286 (62.0%) | 1,320 (53.7%) | 351 (56.8%) | 1,346 (53.5%) |
| Male | 175 (38.0%) | 1,137 (46.3%) | 267 (43.2%) | 1,170 (46.5%) |
| Age |  |  |  |  |
| Mean (SD) | 51 (14) | 56 (18) | 56 (16) | 59 (18) |
| Number of comorbidities |  |  |  |  |
| 0 | 212 (46.0%) | 946 (38.5%) | 235 (38.0%) | 904 (35.9%) |
| 1 | 146 (31.7%) | 747 (30.4%) | 201 (32.5%) | 794 (31.6%) |
| 2 and more | 103 (22.3%) | 764 (31.1%) | 182 (29.4%) | 818 (32.5%) |
| Charlson comorbidity index score |  |  |  |  |
| 0 - 10 | 346 (75.1%) | 1,579 (64.3%) | 421 (68.1%) | 1,544 (61.4%) |
| 11 - 20 | 86 (18.7%) | 521 (21.2%) | 120 (19.4%) | 591 (23.5%) |
| 21 and above | 29 (6.3%) | 357 (14.5%) | 77 (12.5%) | 381 (15.1%) |
| Clinical subgroup |  |  |  |  |
| Immunosuppressed Conditions | 209 (45.3%) | 1,096 (44.6%) | 290 (46.9%) | 946 (37.6%) |
| Non-Haematological Cancers | 66 (14.3%) | 401 (16.3%) | 68 (11.0%) | 594 (23.6%) |
| Other High-Risk Conditions | 122 (26.5%) | 936 (38.1%) | 210 (34.0%) | 915 (36.4%) |
| Unknown | 64 (13.9%) | 24 (1.0%) | 50 (8.1%) | 61 (2.4%) |
| Deprivation |  |  |  |  |
| Most Deprived - 1 | 79 (17.1%) | 518 (21.1%) | 76 (12.3%) | 453 (18.0%) |
| 2 | 87 (18.9%) | 527 (21.4%) | 110 (17.8%) | 520 (20.7%) |
| 3 | 89 (19.3%) | 495 (20.1%) | 110 (17.8%) | 512 (20.3%) |
| 4 | 93 (20.2%) | 437 (17.8%) | 159 (25.7%) | 513 (20.4%) |
| Least Deprived - 5 | 113 (24.5%) | 480 (19.5%) | 163 (26.4%) | 518 (20.6%) |
| Number of vaccine doses |  |  |  |  |
| 0 | 8 (1.7%) | 114 (4.6%) | 8 (1.3%) | 103 (4.1%) |
| 1 - 3 | 386 (83.7%) | 2,215 (90.2%) | 294 (47.6%) | 1,666 (66.2%) |
| 4 and more | 67 (14.5%) | 128 (5.2%) | 316 (51.1%) | 747 (29.7%) |
| All cause hospitalisation or death within 28 days | 23 (5.0%) | 269 (10.9%) | 30 (4.9%) | 275 (10.9%) |

## Contributors

All authors were involved in the conceptualisation and design of the study. AE, GJ, AA, CQ, LA and RB contributed to the acquisition of data. CQ, LA and RB carried out the statistical analysis. AE, CQ and LA drafted the original version of the manuscript. All authors contributed to the interpretation of the results and critical revision of the manuscript. All authors approved the final manuscript. AE is the guarantor for this study. The corresponding author attests that all listed authors meet authorship criteria and that no others meeting the criteria have been omitted.

## Reporting

We used the Strengthening the Reporting of Observational studies in Epidemiology (STROBE) checklist to guide our transparent reporting of our work. A completed STROBE Statement – Checklist of items that should be included in reports of cohort studies is provided.

## Ethics approval

This project uses anonymised individual-level data sources held within the Trusted Research Environment provided by the SAIL Databank at Swansea University, Swansea, UK. All proposals to use SAIL data are subject to review by the independent Information Governance Review Panel (IGRP). This work was approved under proposal number 0911 after careful considerations by IGRP Panel. Access to data is gained through a privacy-protecting safe haven and remote access system referred to as the SAIL Gateway.

## Acknowledgements

This work uses data provided by patients and collected by the NHS as part of their care and support. We would also like to acknowledge all data providers who make anonymised data available for research.

We wish to acknowledge the collaborative partnership that enabled the acquisition and access to the de-identified data, which led to this output. The collaboration was led by the Swansea University Health Data Research UK team under the direction of the Welsh Government Technical Advisory Cell (TAC) and includes the following groups and organisations: the Secure Anonymised Information Linkage (SAIL) Databank, Administrative Data Research (ADR) Wales, Digital Health and Care Wales (DHCW), Public Health Wales, NHS Shared Services Partnership and the Welsh Ambulance Service Trust (WAST). All research conducted has been completed under the permission and approval of the SAIL independent Information Governance Review Panel (IGRP) project number 0911.

We wish to acknowledge the collaborative partnership between SAIL, local health boards and Digital Health and Care Wales that enabled acquisition and access to the de-identified data, and in particular the role of Lydia Herbert, Teena Grenier, Sarah Gage, Emma Williams, Stuart Rees, Jayne Price, Vicky Thomas, Anne Hinchliffe, and Alana Adams in the development of the de-identified individual-level dataset of patients treated with antiviral and nMAb treatments.

## Patient and Public Involvement

The IGRP independent guidance and advice on Information Governance policies, procedures and processes for SAIL Databank. The Panel reviews all proposals to use SAIL Databank to ensure that they are appropriate and in the public interest, and it comprises representatives from various organisations and sectors including members of the public

